# The role of maternal secretor status and human milk oligosaccharides on early childhood development: A systematic review and meta-analysis

**DOI:** 10.1101/2023.11.03.23298019

**Authors:** Martin M. Mulinge, Hellen K. Abisi, Hellen M. Kabahweza, Lydia Okutoyi, Dalton C. Wamalwa, Ruth W. Nduati

## Abstract

Breast milk is the gold standard of infant nutrition, delivering nutrients and bioactive molecules as needed to support optimal infant growth and neurodevelopment. Increasing evidence links the human milk oligosaccharides (HMOs) to these early childhood development milestones. In this PRISMA-compliant systematic review and meta-analysis, we summarised evidence on HMOs and infant brain development, physical growth, and cognitive development. In addition, we compared HMO concentrations between secretor and non-secretor mothers. Searches in PubMed, Scopus, and Web of Science yielded 245 articles, 27 of which were included in the systematic review and 12 in the meta-analysis. The meta-analysis revealed a substantial between-study heterogeneity, I^2^ = 97.3%. The pooled effect was 0.21 (95% CI, −0.41 - 0.83), p = 0.484, indicating that secretors had higher HMO concentrations, though this difference was not statistically significant. At one month of age, 2’FL, 3FL, and 3’SL play an important role in brain maturation and thus play a critical role in cognitive development. Secretors secrete higher concentrations of 2’FL and 3’SL, explaining the benefits to infants of secretor mothers. Growth velocity was correlated to fucosylated and sialylated HMO concentrations, with lower concentrations linked to stunting. In conclusion, evidence from the systematically reviewed articles indicates that HMOs are important for a child’s early development, but the extent to which they have an impact is dependent on maternal secretor status.

## INTRODUCTION

Breastfeeding has undisputed benefits for all aspects of early childhood growth and development, and it is also known that breastfed infants of secretor mothers benefit more than those breastfed by non-secretor mothers [1,2]. In fact, breastfeeding is the gold standard of infant nutrition and is crucial for early childhood development. Suboptimal breastfeeding can disrupt physical growth and neurodevelopment, potentially leading to life-long consequences [3]. The benefit of human milk is the result of millions of years of evolution, which resulted in a perfect multifunctional fluid composed of macronutrients, micronutrients, bioactive molecules, and microbiota [4]. Among the bioactive molecules are human milk oligosaccharides (HMOs), which are non-digestible glycans that have been shown to play a significant role in the stimulation of microbiota growth [5,6]. It is therefore important to conduct an evidence synthesis from the published literature on the role of HMOs on infant physical growth and brain development to establish their contribution to these early childhood development milestones.

It is now known that HMOs are composed of galactose, glucose, fucose, N-acetylglucosamine, and the sialic acid derivative N-acetyl-neuraminic acid, with twelve possible linkages [7]. They are synthesized by lactocytes or imported into milk from the blood supply of the breasts based on infant needs, with colostrum having the highest concentration (>20 g/L) and mature milk having a lower concentration (5-14g/L) [8,9]. It is important to note that HMO composition varies between mothers and is influenced by secretor status, geographic region, season, maternal diet and BMI, gestational age, and duration of lactation [10]. The genetic variation responsible for differences in HMO profiles and concentrations has been mapped to chromosome 19, with other mutations discovered on chromosomes 4, 8, 10, and 11 [11]. Maternal genetics is important because secretor mothers express the FUT2 gene and thus produce milk with an abundance of α1-2 linked fucose moieties such as 2’FL and LNFP I, whereas non-secretor mothers produce little or no such HMOs [12]. Mothers who are Lewis positive express the FUT3 gene and produce milk containing oligosaccharides with α1-3 and α1-4 linkages to fucose such as 3FL, and LNFP II [2,13].

Differences in HMO production based on secretor status have been linked to stimulation of body growth and brain development in the first six months of life [14–16]. Specific HMOs have been linked to infant growth and anthropometry in both high- and low-income populations [17,18]. Another aspect of child development is brain development, which begins a few weeks after conception and continues until the early postnatal years. HMOs are thought to promote early brain development by acting as prebiotics, which are metabolized by gut microbiota and transported from the gut to the brain via the vagus nerve, where they affect brain functions [19,20]. These brain development functions have been linked to sialic acid, which is abundant in HMOs. Notably, sialic acid is an essential nutrient for brain development because it crosses the blood-brain barrier and reaches the frontal cortex grey matter, where it is involved in ganglioside formation and myelination, both of which are key components of developing cortical grey matter and white matter [21]. Therefore, it is critical to identify HMOs that are important in infant growth and prioritize the synthetic HMOs that can be included in formulas cognizant of the fact that some HMOs may increase or mitigate the risk of excessive weight gain and obesity in childhood which persists in adulthood. As a result, it is critical to identify HMOs that are important in infant growth and neurodevelopment, given that technology exists to synthesize HMOs, which can be used as a supplement on infants who receive inadequate breastmilk.

A review of published systematic reviews up to June 2023 yielded 5 articles relevant to this topic. Two studies explored the effect of breastfeeding on infant body composition [22,23], while three articles focused on the role of HMOs on neurodevelopment and cognitive functions in children [24–26]. All of these systematic reviews address the effect of human milk components on either infant growth or neurodevelopment, but none have systematically evaluated and synthesized evidence on the effect of HMOs on both infant growth and cognitive development, which are two intertwined concepts of early human development. The primary goal of this systematic review is to summarize evidence on the influence of HMOs on brain development, infant growth velocity and cognitive development using Preferred Reporting Items for Systematic Reviews and Meta-analyses (PRISMA) guidelines. The secondary goal was to perform meta-analyses on HMO concentration differences between secretors and non-secretors, as well as to determine between-study heterogeneity.

## METHODS

The PRISMA guidelines were followed when conducting this systematic review and meta-analysis [27]. An ethical review was not necessary because this was a review of studies that had already been published.

### Data sources and search strategy

The literature search was conducted by HK. Articles published in English from three databases (PubMed, Scopus and Web of Science) using identified keywords and index terms. The search terms were divided into three concepts, combined with the boolean phrases “AND” between concepts and “OR” within concepts: [1] breastfeeding AND [2] human milk oligosaccharides OR human milk sugars AND [3] infant growth OR brain development OR neurodevelopment OR cognitive development. The search was limited to articles published from 2000 informed by an extensive review published Kunz and colleagues focusing on the structural, functional and metabolic aspects of HMOs [8], up to June 30, 2023. We conducted the search and limited the selection to original articles published in English, excluding grey literature, using the syntax shown in supplementary table 1. The search terms used on PubMed were modified for Scopus and Web of Science database searches.

### Eligibility criteria

We developed a Population, Intervention, Comparators, Outcome, and Study design approach as the eligibility criteria, determined by the following research question: What effect do HMOs in human milk [Intervention] have on brain development, infant growth velocity, and neurodevelopment [outcomes] in breastfed infants [population]? Population: studies that included nursing mothers and their infants were eligible. Intervention: exposure to HMOs via breastfeeding. Comparators: Secretors vs non-secretor mothers. Outcome: brain development, infant growth velocity and neurodevelopment. Study design: observational and intervention studies. Exclusion: Inappropriate study types (animal studies, in vitro studies, abstracts from conferences, reviews, and meta-analyses), as well as articles that did not link HMOs to brain development, infant growth rate, or cognitive development. Two authors (MM and HA) conducted the literature selection for eligibility by removing all undoubtedly irrelevant articles before using rayyan (http://rayyan.qcri.org), a web tool that speeds up the screening process by multiple authors concurrently [28].

### Data extraction

The data extracted from each article included: author, year of publication, country, study design, population size (n), milk samples (n), HMO profiled (n), HMO profiling technique, proportion of secretors (%) and non-secretors mothers (%), and effect of HMOs on brain development, growth velocity and cognitive development. MM & HA independently extracted data into Microsoft excel template. When there were disagreements, a consensus was reached after discussing with LO.

### Quality Assessment

The study’s quality was independently assessed by MM, HA, and HK. A majority of the studies were observational and therefore Newcastle-Ottawa Scale (NOS) was used to assess the risk of bias [29]. The NOS is a three-domain scale that assesses participant selection, comparability, and outcome. Each study may receive up to four points in the selection section (if the study is truly representative of the community and prospective) and three points in the outcome section (if outcome measures data are collected as accurately as possible and the cohort follow-up rate is >80%). Depending on the adjustment for confounders, a maximum of two points can be awarded for the comparability section. As a result, a study can be graded from 0 to 9. A high-quality study will receive 8-9 points, a medium-quality study will receive 6-7 points, and a low-quality study will receive 6 points. Cross-sectional studies receive up to 6 stars, while longitudinal studies receive up to 9 stars.

### Data analysis

The primary goal of all of the studies was to determine the effect of breastmilk HMOs on brain development, infant growth velocity, or cognitive development of breastfed infants. If means and standard deviations (SD) for HMO concentrations were not reported, they were calculated from the median and (IQR) using an online calculator https://www.math.hkbu.edu.hk/~tongt/papers/median2mean.html. A weighted mean was computed using the Hmisc R package to determine the proportion of secretor versus non-secretor mothers. To compare concentrations between secretor and non-secretor mothers, meta-analyses were performed using the meta R package. The majority of studies were cross-sectional, but if longitudinal, the earliest timepoint was used. The standardized mean difference (SMD) was used as the outcome measure to assess statistical heterogeneity in the studies that were included in the meta-analysis. Since we expected significant between-study heterogeneity, we used a random-effects model to pool effect sizes. The heterogeneity variance τ^2^ was estimated using the restricted maximum likelihood (REML) estimator. Because percentages are easier to report, the I^2^ statistic is reported in addition to the τ^2^ statistic. The confidence interval around the pooled effect was computed using Knapp-Hartung adjustments. Studentized residuals and Cook’s distances were used to identify whether studies were outliers and/or influential in the context of the model. Publication bias was assessed through the Egger weighted regression and visualized using funnel plots.

## RESULTS

### Eligible studies

A PRISMA flowchart of the study selection process is shown in Figure 1. HK conducted an initial search in the three databases, and the results were 245 articles. After the duplicates were removed, 176 articles were left. After evaluating titles and abstracts for study type, or irrelevance, a further 61 articles were eliminated. MM and HK downloaded the full texts of the remaining 69 articles and reviewed them for eligibility. At this point, 42 articles were removed from consideration because they did not meet the inclusion criteria, leaving only 27 articles in the systematic review: [17,18,38–47,30,48–54,31–37]. Additionally, 12 studies that compared HMO concentrations in secretors and non-secretor mothers were included in the meta-analysis [30,31,42,43,48,33–36,38–41].

**Figure 1.**
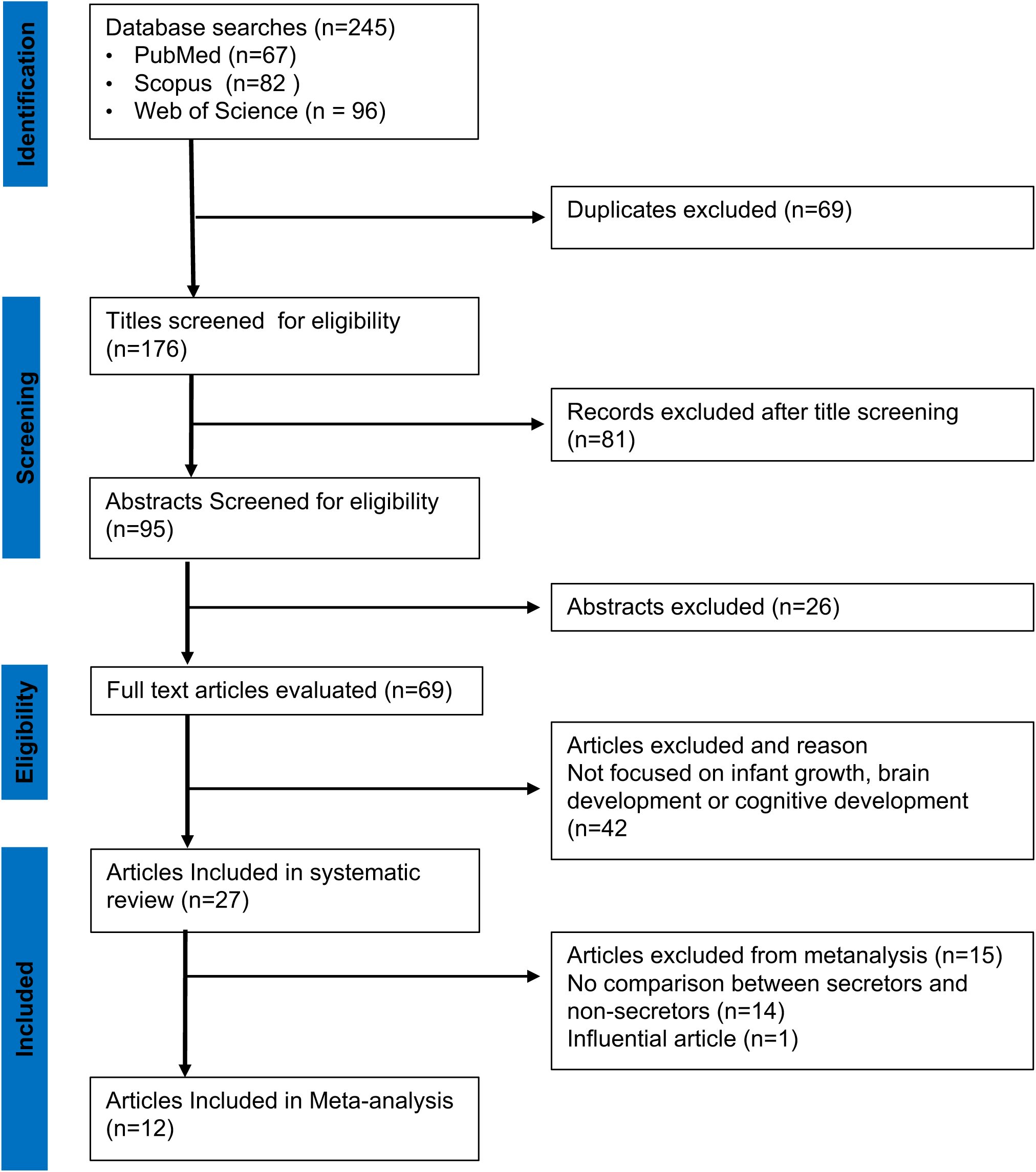
PRISMA Diagram of Study Selection. The relevant number of articles at each step are indicated.

### Quality assessment

According to the NOS findings (Supplemental Table 2), 17 studies (71%) had medium quality, which was attributed to higher scores in comparability and outcome aspects. The main reason for the lower-quality studies was a lack of detail about potential baseline differences between groups or confounding variables. The aspect of outcome (HMO profile) within the NOS was adequately conducted in most studies in accordance with the study’s research question.

### Study characteristics

Table 1 summarizes the sociodemographic and HMO data from the 27 studies that were included in this review. The selected studies were conducted in 21 countries: Australia [31,43]; Bangladesh [53]; Brazil [36,50]; China [32,41]; Denmark [54]; Finland [30]; France [48]; Gambia [18]; Germany [39]; Kenya [42]; Malawi [47,51]; Netherlands [44]; Philippines [52]; Singapore [33]; Spain [40]; Sweden [49]; and USA [17,34,35,37,38,45]. In addition, there was one European Union study involving milk samples collected from France, Italy, Norway, Portugal, Romania, Spain and Sweden [46]. A total of 7,648 milk samples were examined in the 27 studies, which involved 4,066 mothers and 4,103 infants. The differences between mothers and infants are due to twin pregnancies. The weighted mean of secretor mothers was 77.8% (95% CI, 60.3-95.3).

**Table 1:**
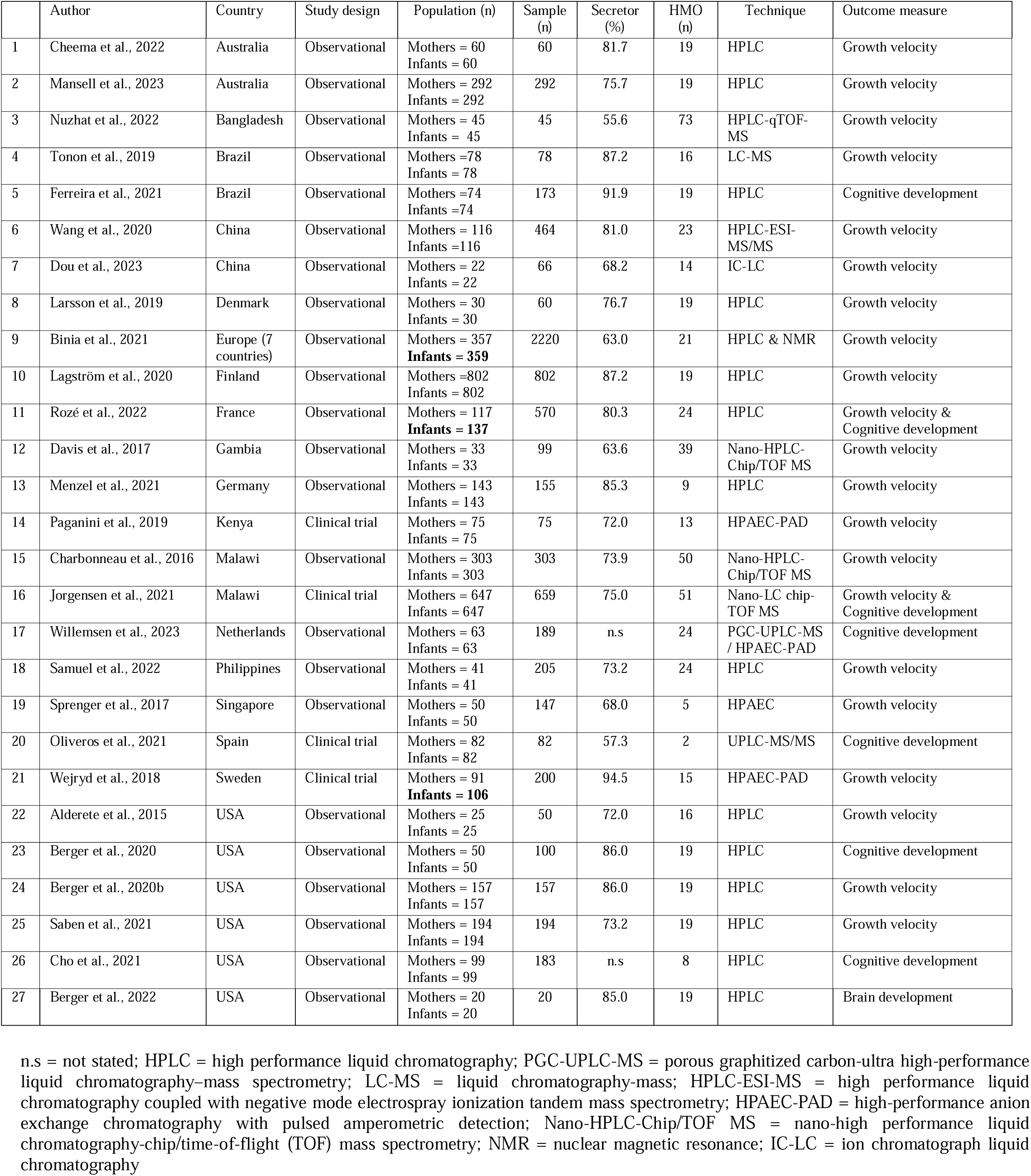
Summary of selected studies (n = 27).

|  | Author | Country | Study design | Population (n) | Sample (n) | Secreter (%) | HMO (n) | Technique | Outcome measure |
| --- | --- | --- | --- | --- | --- | --- | --- | --- | --- |
| 1 | Cheema et al., 2022 | Australia | Observational | Mothers = 60<br>Infants = 60 | 60 | 81.7 | 19 | HPLC | Growth velocity |
| 2 | Mansell et al., 2023 | Australia | Observational | Mothers = 292<br>Infants = 292 | 292 | 75.7 | 19 | HPLC | Growth velocity |
| 3 | Nuzhat et al., 2022 | Bangladesh | Observational | Mothers = 45<br>Infants = 45 | 45 | 55.6 | 73 | HPLC-qTOF-MS | Growth velocity |
| 4 | Tonon et al., 2019 | Brazil | Observational | Mothers = 78<br>Infants = 78 | 78 | 87.2 | 16 | LC-MS | Growth velocity |
| 5 | Ferreira et al., 2021 | Brazil | Observational | Mothers = 74<br>Infants = 74 | 173 | 91.9 | 19 | HPLC | Cognitive development |
| 6 | Wang et al., 2020 | China | Observational | Mothers = 116<br>Infants = 116 | 464 | 81.0 | 23 | HPLC-ESI-MS/MS | Growth velocity |
| 7 | Dou et al., 2023 | China | Observational | Mothers = 22<br>Infants = 22 | 66 | 68.2 | 14 | IC-LC | Growth velocity |
| 8 | Larsson et al., 2019 | Denmark | Observational | Mothers = 30<br>Infants = 30 | 60 | 76.7 | 19 | HPLC | Growth velocity |
| 9 | Binia et al., 2021 | Europe (7 countries) | Observational | Mothers = 357<br>**Infants = 359** | 2220 | 63.0 | 21 | HPLC & NMR | Growth velocity |
| 10 | Lagström et al., 2020 | Finland | Observational | Mothers = 802<br>Infants = 802 | 802 | 87.2 | 19 | HPLC | Growth velocity |
| 11 | Rozé et al., 2022 | France | Observational | Mothers = 117<br>**Infants = 137** | 570 | 80.3 | 24 | HPLC | Growth velocity & Cognitive development |
| 12 | Davis et al., 2017 | Gambia | Observational | Mothers = 33<br>Infants = 33 | 99 | 63.6 | 39 | Nano-HPLC-Chip/TOF MS | Growth velocity |
| 13 | Menzel et al., 2021 | Germany | Observational | Mothers = 143<br>Infants = 143 | 155 | 85.3 | 9 | HPLC | Growth velocity |
| 14 | Paganini et al., 2019 | Kenya | Clinical trial | Mothers = 75<br>Infants = 75 | 75 | 72.0 | 13 | HPAEC-PAD | Growth velocity |
| 15 | Charbonneau et al., 2016 | Malawi | Observational | Mothers = 303<br>Infants = 303 | 303 | 73.9 | 50 | Nano-HPLC-Chip/TOF MS | Growth velocity |
| 16 | Jorgensen et al., 2021 | Malawi | Clinical trial | Mothers = 647<br>Infants = 647 | 659 | 75.0 | 51 | Nano-LC chip-TOF MS | Growth velocity & Cognitive development |
| 17 | Willemsen et al., 2023 | Netherlands | Observational | Mothers = 63<br>Infants = 63 | 189 | n.s | 24 | PGC-UPLC-MS / HPAEC-PAD | Cognitive development |
| 18 | Samuel et al., 2022 | Philippines | Observational | Mothers = 41<br>Infants = 41 | 205 | 73.2 | 24 | HPLC | Growth velocity |
| 19 | Sprenger et al., 2017 | Singapore | Observational | Mothers = 50<br>Infants = 50 | 147 | 68.0 | 5 | HPAEC | Growth velocity |
| 20 | Oliveros et al., 2021 | Spain | Clinical trial | Mothers = 82<br>Infants = 82 | 82 | 57.3 | 2 | UPLC-MS/MS | Cognitive development |
| 21 | Wejryd et al., 2018 | Sweden | Clinical trial | Mothers = 91<br>**Infants = 106** | 200 | 94.5 | 15 | HPAEC-PAD | Growth velocity |
| 22 | Alderete et al., 2015 | USA | Observational | Mothers = 25<br>Infants = 25 | 50 | 72.0 | 16 | HPLC | Growth velocity |
| 23 | Berger et al., 2020 | USA | Observational | Mothers = 50<br>Infants = 50 | 100 | 86.0 | 19 | HPLC | Cognitive development |
| 24 | Berger et al., 2020b | USA | Observational | Mothers = 157<br>Infants = 157 | 157 | 86.0 | 19 | HPLC | Growth velocity |
| 25 | Saben et al., 2021 | USA | Observational | Mothers = 194<br>Infants = 194 | 194 | 73.2 | 19 | HPLC | Growth velocity |
| 26 | Cho et al., 2021 | USA | Observational | Mothers = 99<br>Infants = 99 | 183 | n.s | 8 | HPLC | Cognitive development |
| 27 | Berger et al., 2022 | USA | Observational | Mothers = 20<br>Infants = 20 | 20 | 85.0 | 19 | HPLC | Brain development |

n.s = not stated; HPLC = high performance liquid chromatography; PGC-UPLC-MS = porous graphitized carbon-ultra high-performance liquid chromatography–mass spectrometry; LC-MS = liquid chromatography-mass; HPLC-ESI-MS = high performance liquid chromatography coupled with negative mode electrospray ionization tandem mass spectrometry; HPAEC-PAD = high-performance anion exchange chromatography with pulsed amperometric detection; Nano-HPLC-Chip/TOF MS = nano-high performance liquid chromatography-chip/time-of-flight (TOF) mass spectrometry; NMR = nuclear magnetic resonance; IC-LC = ion chromatograph liquid chromatography

### Pooled effect and Heterogeneity of the reported results

Twelve articles comparing HMO concentrations in secretor and non-secretor mothers were included in the meta-analysis. Overall, the pooled Hedges’ g (bias corrected standardised mean difference) was 0.21 (95% CI, −0.41-0.83), with the majority of estimates being positive. In the context of our meta-analysis, this indicates a small effect, with secretors having higher HMO concentrations than non-secretors, but the difference between the two groups is not statistically significant, p = 0.48. Moreover, the 95% prediction interval ranged from g = −2.47 to 2.89, indicating that, although the average outcome is estimated to be positive, in some studies the true outcome may in fact be negative. The REML method estimated a substantial between-study heterogeneity variance (I^2^ value of 97.3% (95% CI, 96.6-97.9), p <.001. Looking at τ^2^ = 1.54 (95%CI, 0.85-3.62), p <.001, we can see that the confidence interval does not include zero, indicating that the variance of true effect sizes is significantly greater than zero (Figure 2). According to Higgins and Thompson’s I^2^ statistic, 15/19 HMOs profiled were generated by studies with moderate to substantial heterogeneity. The τ^2^ statistic included a zero in the remaining HMOs, indicating homogeneity (Table 2). The g of 2’FL produced exclusively by secretors was 2.8 (95% CI, 2.40-3.20), p <.001. Other key HMOs with significantly higher bias corrected SMDs in secretors were LNFP-1, 1.45 (95% CI, 1.08-1.82), p <.001, and DFlac, 2.78 (95% CI, 0.63-4.97) p = 0.024. Non-secretor mothers had significantly higher concentrations of the following HMOs compared to secretor mothers: LNFP II, −1.97 (95% CI, to −2.92 to −1.02), p = <.001 and FDSLNH, −2.07, (95% CI, −2.58 to −1.57), p <.001. The concentrations of three key HMOs involved in early childhood development (3’SL, 3FL, and 6’SL) are highlighted. 3’SL concentrations were higher in secretor mothers, but the averaged bias corrected SMD did not differ significantly from zero, 0.21, (95% CI, −0.17-0.59), p = 0.23, while 3FL and 6’SL concentrations were higher in secretors and non-secretors, respectively (Table 2). An examination of the studentized residuals revealed that one study [38], was a significant outlier that influenced the pooled statistic. Although the mean was not particularly extreme, the narrow standard deviations influenced its weight on the overall pooled effect. When it was removed from the analysis of studies profiling 2’FL and 3’SL, the value of 2 decreased from 13.8 to 0.3 for 2’FL and from 10.6 to 0.19 for 3’SL. The pooled effect of g = 2.8 was slightly lower than our initial estimate of g = 3.8 for 2’FL and 0.21 from 1.21 for 3’SL, but it is still within the same order of magnitude (Supplementary Table 3). There was no evidence of outlier HMOs in the final fitted model. Eggers’ test did not indicate the presence of funnel plot asymmetry: intercept 2.29 (95% CI, −3.49-8.07), t = 0.78, p = 0.44, which is consistent with funnel plot (Figure 3).

**Figure 2.**
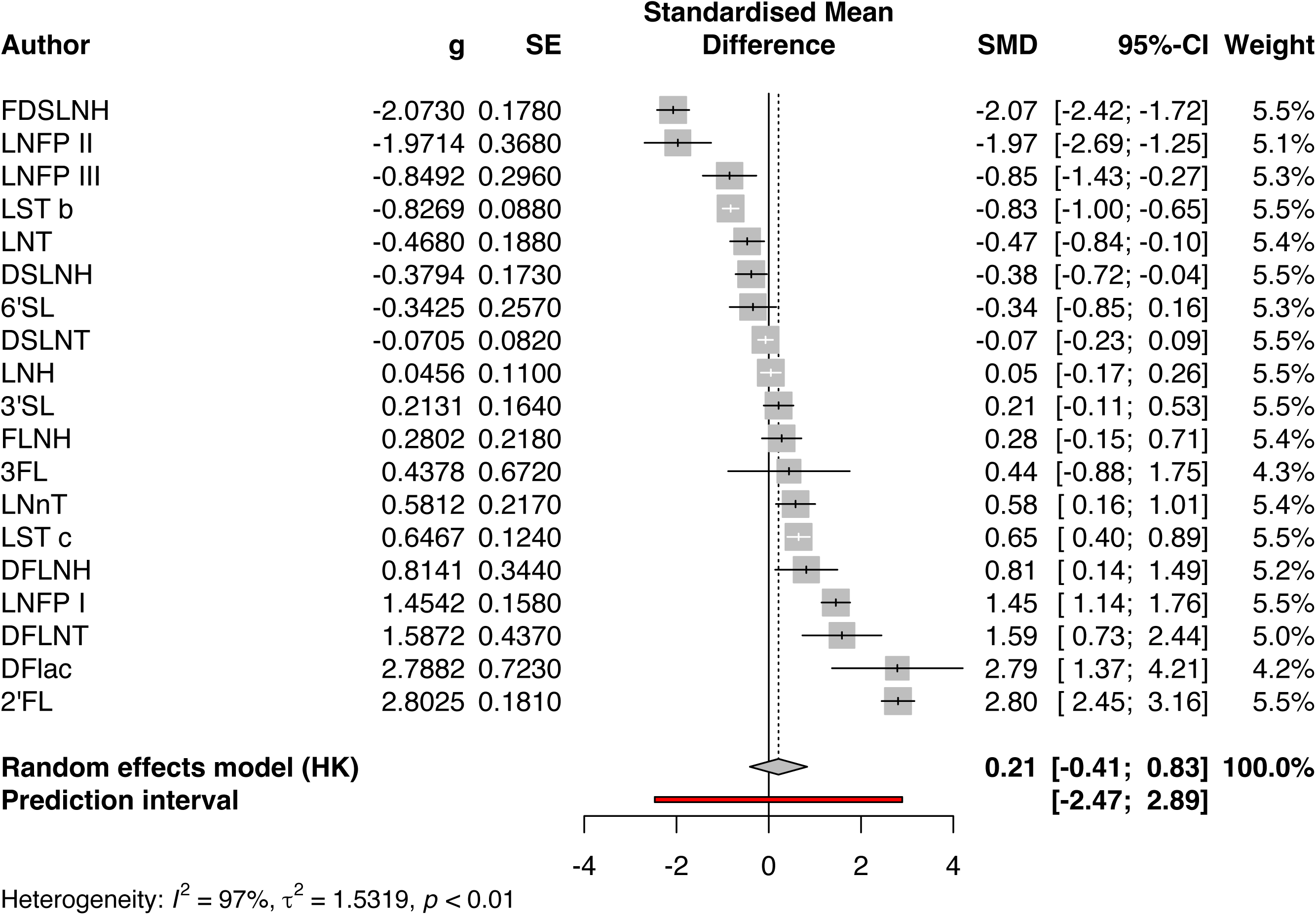
Forest plot of the standardised mean difference (g) of HMO concentration between secretors and non-secretors. A positive effect (g) indicates a higher concentration in secretors while a negative effect (-g) indicates higher HMO concentration in non-secretors. Note that the Confidence Intervals in this figure differ from those in Table 2 due to Hartung-Knapp (HK) adjustment for random effects model (df = 18). Prediction interval is based on t-distribution (df = 17). g = Hedges’ g (bias corrected standardised mean difference). SMD = standardised mean difference, SE = standard error, 2’FL = 2-fucosyllactose, 3FL = 3-fucosyllactose, DFLac = difucosyllactose, DFLNH = difucosyllacto-N-hexaose, DFLNT = difucosyllacto-N-tetraose, FLNH = fucosyllacto-N-hexaose, LNFP I = lacto-N-fucopentaose I, LNFP II = lacto-N-fucopentaose II, LNFP III = lacto-N-fucopentaose III, 3’SL = 3-sialyllactose, 6’SL = 6-sialyllactose, DSLNT = disialyllacto-N-tetraose, DSLNH = disialyllacto-N-hexaose, LST b = sialyllacto-N-tetraose b, LST c = sialyllacto-N-tetraose c, FDSLNH = fucodisialyllacto-N-hexaose, LNH = lacto-N-hexaose, LNnT = lacto-N-neotetraose, LNT = lacto-N-tetraose.

**Figure 3.**
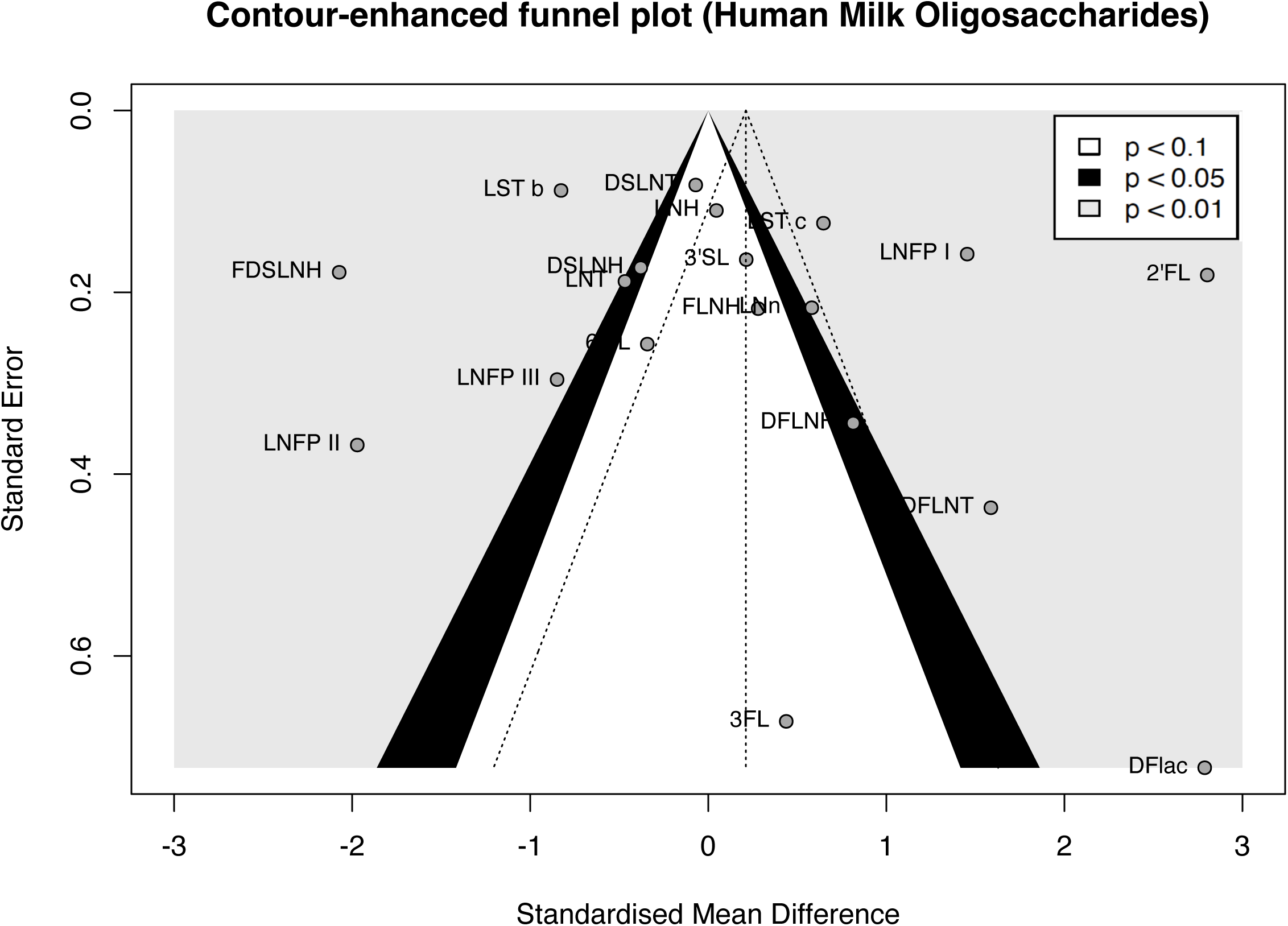
Contour enhanced funnel plot for 19 HMOs concentrations between secretors and non-secretors. It shows a scatter plot of the studies’ observed effect sizes on the x-axis against a measure of their standard error on the y-axis. The higher values on the y-axis represent HMOs with the lower standard errors. In this diagram there is no indication of asymmetry. 9 HMOs are found in regions of high significance (p <0.01) in [grey zone], 3 in regions of significance (p 0.05) in [black zone], and 6 HMOs in regions of no significance (p < 0.1) in [white zone].

**Table 2:**
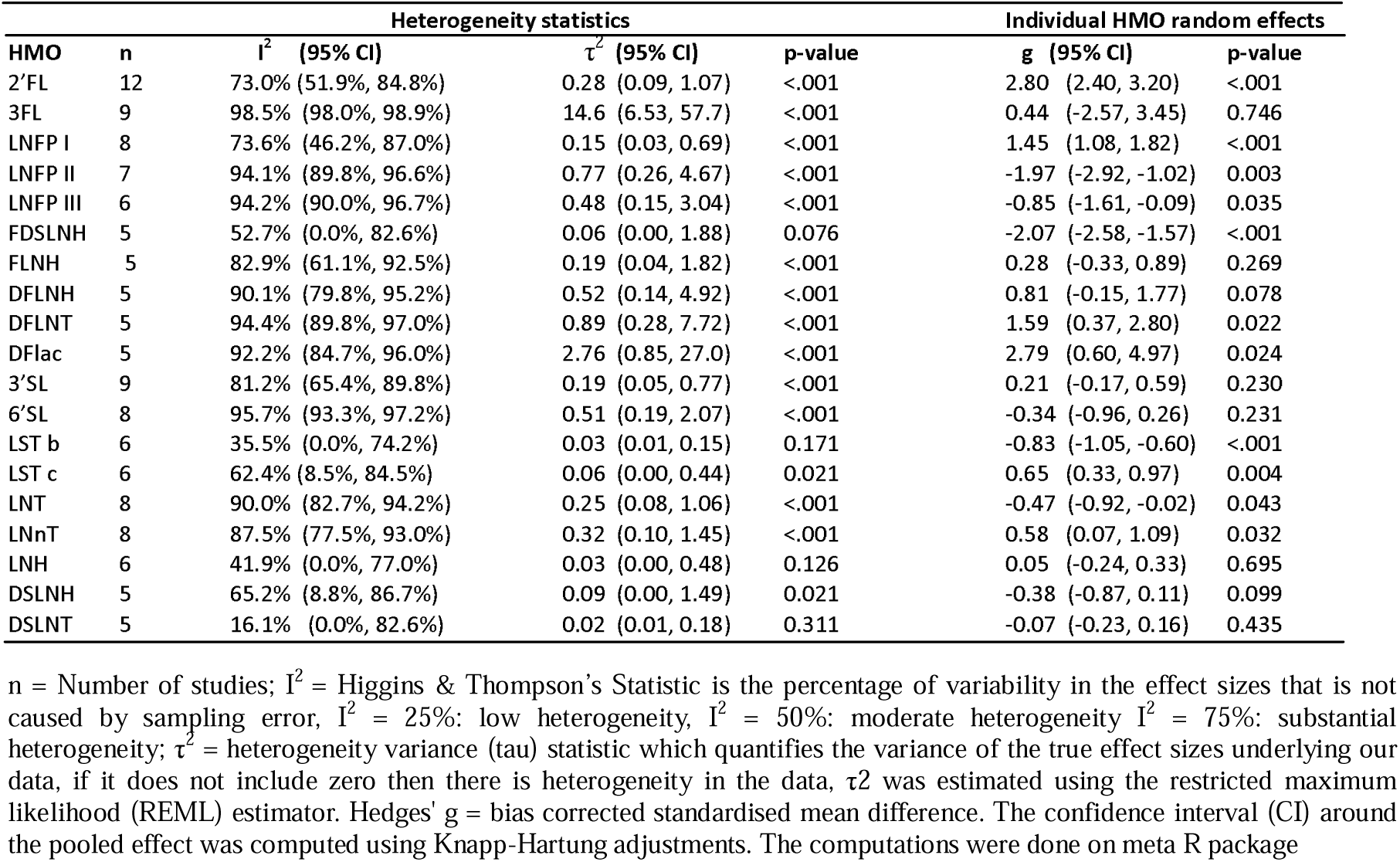
The heterogeneity and estimated average standardized mean differences of 19 HMOs computed by random-effects model.

### Analytical methods for characterizing HMOs

Various analytical techniques were used to identify HMOs, depending on the researchers’ goals. The majority of the studies (14/27) used high-performance liquid chromatography (HPLC). 4/27 studies used high-pH anion exchange chromatography (HPAEC), while 7/27 used mass spectrometry (MS) as a detector. The following 19 HMOs were studied in multiple studies and were included in the meta-analysis: fucosylated HMOs [2-fucosyllactose (2’FL), 3-fucosyllactose (3FL), difucosyllactose (DFLac), difucosyllacto-N-hexaose (DFLNH), difucosyllacto-N-tetraose (DFLNT), fucosyllacto-N-hexaose (FLNH), lacto-N-fucopentaose I (LNFP I), lacto-N-fucopentaose II (LNFP II), lacto-N-fucopentaose III (LNFP III)]; sialylated HMOs [3-sialyllactose (3’SL), 6-sialyllactose (6’SL), disialyllacto-N-tetraose (DSLNT), disialyllacto-N-hexaose (DSLNH), sialyllacto-N-tetraose b (LST b), sialyllacto-N-tetraose c (LST c)]; fucosylated and sialylated HMOs [fucodisialyllacto-N-hexaose (FDSLNH)]; and undecorated HMOs [lacto-N-hexaose (LNH), lacto-N-neotetraose (LNnT), lacto-N-tetraose (LNT)]. Given that different authors used different abbreviations in their published articles, we used Lars Bode’s (2012) abbreviations for consistency in our manuscript [6].

### HMOs involved in brain tissue organization

The authors used a 3.0-Tesla MRI scanner to correlate MRI indices of tissue microstructure and regional cerebral blood flow (rCBF) in infants with HMO concentrations. Most notably in the cortical mantle, 2’FL was connected to decreased fractional anisotropy (FA), increased mean diffusivity (MD), and decreased rCBF values, suggesting that this HMO is crucial for the formation of dendritic arbours and synapses for circuit formation. In the developing white matter, 3FL and 3’SL were linked to higher FA, lower MD, and higher rCBF values, indicating that these HMOs may improve structural connectivity in the brain. While 2’FL, 3FL, and 3’SL were linked to early brain maturation, 6’SL was not associated to these MRI indices [37].

### HMOs involved in cognitive development

This section summarizes the involvement of HMOs in cognitive development in six studies using various techniques such as: Ratings of Everyday Executive Functioning (REEF); Behaviour Rating Inventory of Executive Function (BRIEF); Age and Stages Questionnaire (ASQ); Bayler III Cognitive Scale; and Mullen Scales of Early Learning (MSEL). The REEF and BRIEF scores were used to determine executive functions at different time points. 2’FL and total fucosylated HMOs were associated with better executive functions at 3 years, and 2’FL was associated with 4-behavioral tasks at 2 and 12 weeks, regardless of whether the infants were exclusively breastfed or partially breastfed; however, 3’SL, 6’SL, and total Sialylated HMOs were associated with worse executive functions at 3 years [44]. The ASQ score was used in 2/6 studies to assess the development of communication skills, fine motor skills, gross motor skills, and personal social skills. Positive associations were determined for: 3FL for communication skills; DSLNT, FLNH for fine motor skills; and negative associations for: 2’FL, DFLac, LNH, and 3’SL for personal social skills, LNnT for problem solving, and LNFP III for gross motor skills [50]. In a study by Roze and colleagues, LNFP III was associated with ASQ score in secretor mothers but not in the entire cohort [48]. The Bayler III cognitive scale, which measures cognitive development at 24 months using five domains: cognitive, language, motor, adaptive, behavior, and social-emotional development, was used in 2/6 studies. 2’FL, LNH, and FLNH were positively associated with cognitive development, while LSTb was negatively associated with cognitive development [34], whereas 2’FL and 6’SL were positively associated with cognitive development in secretors [40]. The MSEL is a tool for assessing infant cognitive development that is divided into five subdomains: fine motor, gross motor, visual reception, receptive language, and expressive language. In A-tetra+ HMO mothers, there was a positive relationship between 3’SL and early learning composite, receptive language t score [45].

### HMOs involved in infant growth velocity

Various anthropometrics indexes were used to report infant growth in 19 studies. Weight gain was reported as weight z score or weight for age z score in 10/19 studies. The following studies found positive associations between HMOs and weight gain: 2’FL [30,43,54], 3FL [30,38,43], 3’SL [18,30,38,43], LNnT [35], LNFP II [38], DFLac [30,43,54], LST b [38], LNH [41], DFLNH [43], DSLNH [38], DFLNT [43], and negative associations for 3’SL [31,32], 6’SL [43], LNnT [30,32,54], LNFP II [35], LNH [31], LNFP I [17], LST b [30], LST c [18], DFLNH [54], DSLNT [32,35], FDSLNH [31].

The second metric, height gain, was reported in 12/19 studies as either weight for length z score, length for age z score, or height for age z score. The following HMOs had a positive correlation with height gain: 2’FL [30,31,47,48], 3FL [43], 3’SL [39,43], LNnT [39], LST-a [49,51], DFLNH [41,43,51], and DFLNT [43] while a negative correlation was linked with: 3FL [52], 3’SL [46], 6’SL [49], LNnT [30,39,43,46], LNT [31,39], LNFP I [39,47], LNFP II [32], LST b [30,31,49], and FLNH [39].

The third infant growth metric was head growth which was reported in 7/19 studies as either head circumference for age z score or head circumference z score. This metric which also serves as an index of cognitive development and was positively correlated with the following HMOs: 2’FL [31], 3’SL [51,54], 6’SL [48], LNFP III [46], LST-a [49], DFLac [54], and negatively correlated with 6’SL [49,51], LNFP III [18], LNFP I [18], DFLNH [54], LST b [49], and DSLNH [31].

Body mass index, a gauge of body composition, was reported as body mass index for age z score in 7/19 studies. The following were reported to have positive associations with this metric: DFLac [43], LST b [43], and DFLNT [43], while a negative association was reported for 2’FL [39], 6’SL [54], LNnT [30], and LNnT [39]. The percentage of fat mass change was independently reported in 4/19 studies and is closely related to body mass index and weight gain. Positive correlations for were noted for: 2’FL [43], 3’SL [38], 6’SL [38], LNFP III [38], LNFP II [17,38], DSLNT [17,38], LST b [38], FDSLNH [17], DSLNH [38], DFLNT [43], while negative association was reported for: 6’SL [43], LNnT [17,54], LNFP I [17], LNFP III [43], and DFLNH [54]. In 4/19 studies, the change in fat-free (lean) mass was reported, and it was positively related to 3FL [43], 3’SL [43,48], DFLac [43], DFLNH [43] and DFLNT [43], whereas a negative correlation was found for: LNT [48], LNFP I [17], LST c [48] [49]. Skinfold thickness z-score, a novel metric, was positively correlated with 3FL and 3’SL and negatively correlated with 6’SL, LNnT, and LST c [31].

### HMOs predominant in stunting or severe acute malnutrition

Two studies focused on the effects of undernutrition on HMO composition and consequently on stunting [47], and severe acute malnutrition (SAM) [53], in Malawi and Bangladeshi children respectively. In these studies, ultrasensitive qTOF-MS was used to conduct robust HMO profiling, yielding 50 and 73 HMOs, respectively. Lower levels of fucosylated and sialylated HMO were linked to stunting in Malawian infants [47]. In contrast, total sialylated HMOs were linked to a higher risk of SAM in Bangladeshi children, whilst there was no evidence of a significant association between SAM and fucosylated or undecorated HMOs [53].

## DISCUSSION

Our study provides a comprehensive analysis of 19 HMOs on early childhood milestones based on published studies in order to better understand their roles, which have remained understudied when compared to other human milk components. We collected and pooled data that suggests that 2’FL, 3FL, and 3’SL were associated with early brain maturation, which is important in understanding the optimal growth and cognitive development benefits observed in breastfed infants and superior outcomes on infants nursed by secretor mothers [37,55]. Suboptimal breastfeeding or poor maternal nutrition resulted in lower total fucosylated and sialylated HMOs, resulting in stunting. Overnutrition has been linked to excessive weight gain in children in some studies when breastfeeding children are also supplemented with formulas containing 2’FL. Interestingly, LNFP II was found to be protective against pediatric obesity, and early supplementation with this HMO may help to prevent childhood obesity, which can persist into adolescence and adulthood. The pooled studies were highly heterogeneous, with secretors having higher HMO concentrations despite the fact that the difference was not statistically significant. Nevertheless, key HMOs involved in early childhood development, such as 2’FL and 3’SL, were predominant in secretors explaining the benefits to their nursing infants.

Among the 224 identified HMOs [56], this review focused on 19 well-profiled HMOs and their effects on brain development, infant growth velocity, and neurodevelopment. As expected, there was evidence of between-study heterogeneity, which ranged from low to substantial in accordance with Higgins and Thompson’s I^2^ statistic [57]. In this study, the pooled concentrations of 2’FL, DFlac, DFLNT, LNFP I, LST c, LNnT, FDSLNH, LNFP II, LNFP III, LST b, and LNT were statistically different between secretors and non-secretors, consistent with previous studies that found HMO concentrations varied by maternal secretor status [58–61]. The selected studies also confirmed the previously reported observation that secretor status varies by geographical location, with the highest proportions of secretor mothers in South America and Northern Europe and the lowest proportions in Africa and Asia [62–67]. The expression of the FUT2 gene, whose locus is mapped to chromosome 19, is now known to be the primary driver of this genetic variation[11]. A compensatory mechanism occurs via expression of FUT3 gene (Lewis positive) where mothers produce milk containing oligosaccharides with α1-3 and α1-4 linked fucoses including 3FL, and LNFP II [2,13]. This finding confirms that evolutionary dynamics ensured that non-secretors retained the ability to produce key fucosylated HMOs required for early childhood development though not as efficiently as secretors. Seasonality was identified as a key environmental factor regulating HMO synthesis in Gambian mothers [18], and similar patterns were observed in Israeli mothers, with HMO concentrations significantly lower in the summer [66]. Given that seasons are a common phenomenon all over the world, this is a critical factor that should be considered in study designs. Though not frequently reported as a probable causal factor of interest, infants’ biological sex influenced HMO in Brazilian mothers [36], which is consistent with the findings of Asher and colleagues in a study in which seasonality was adjusted for [66]. The milk collection technique, which was dependent on geographical location, could also have contributed to the observed differences. Moossavi and colleagues discovered that using breast pumps was associated with lower relative abundance of *Bifidobacterium spp.* and higher levels of DSLNH, emphasising the indirect role of the microbiome, which will be discussed in detail under infant growth [68]. Maternal weight and BMI adjusted for in 13 studies may have also played a role in HMO concentrations, consistent with previous findings [64,69].

Breastfeeding that starts as soon as the baby is born and continues exclusively for 6 months and extended breastfeeding for up to 2 years have been linked to higher cognitive functions in children studied using REEF, BRIEF, ASQ and MSEL tools since the early ages[24,70]. This has been attributed to sialic acid which is abundant in sialylated HMOs and its role in in brain development, neuronal transmission, and synaptogenesis which is well established [71,72]. This link was clarified by Berger and colleagues’ groundbreaking study [37], which demonstrated the link between key fucosylated and sialylated HMOs on brain maturation, explaining why breastfed infants of secretor mothers benefit from more health benefits than those of non-secretor mothers. It also suggests that in the absence of 2’FL supplementation, infants of non-secretor mothers who express no 2’FL and less 3’SL benefit less, or that compensation occurs through unknown mechanisms. With the link between HMOs and brain development and thus higher cognitive functions established, we now have a solid foundation to advocate for the importance of exclusive breastfeeding after birth to reap the greatest benefits, as the benefits identified at one month diminish after 6 months indicating that early exposure is temporal and the most critical time which has an influence on infant cognitive development [34]. This knowledge may also benefit preterm infants born when their mothers’ entero-mammary pathway is not fully developed [73], by exposing them to mature donor milk from confirmed secretor mothers which is loaded with sufficient 2’FL and microbiome for optimal development. In a study of preterm infants, LNFP III was significantly associated with higher cognitive development in infants born to Secretor(+) Lewis(+) mothers [48], indicating that it could play a significant role in this vulnerable population but only in secretor mothers’ infants. The microbiota is also involved in cognitive development via the gut-brain axis. For instance, the relative abundances of *Veillonella spp.*, a critical microbiota regulating γ-aminobutyric acid (GABA), a critical neurotransmitter for early brain development [74], were inversely correlated with concentrations of sialylated HMOs [63]. Some HMOs, including LNT and DSLNT, were linked to poor cognitive development and were notably higher in non-secretor mothers in our pooled analyses [34,50].

While the link between cognitive development and sialylated HMOs is well established [37], the connection between HMOs and growth velocity is blurred and indirect given that growth functions are largely influenced by lactose lipids, lactose, and proteins [75,76]. The relatively low protein and lipid concentrations in human milk, but higher HMO concentrations, are an adaptation to the lower needs of infant growth but optimal development of higher order cognitive functions at a younger age than non-human mammals who gain weight at a much faster rate [56,77]. Nonetheless, the importance of HMOs in physical growth can be seen in the context of malnutrition, where breastmilk from mothers with stunted children contained lower levels of total HMOs than breastmilk from mothers with healthy infants, with adverse effects for infants of non-secretor mothers who are unable to compensate for deficiencies in fucosylated HMOs through increased output of other (e.g., sialylated) HMOs, resulting in milk that does not support infant growth [47]. Addressing undernutrition is a global issue because, as of 2020, 144 million children under the age of five were stunted, which disadvantages them mainly because they have difficulty learning in school, earn less as adults, and face barriers to participation in their communities [78]. 2’FL, a critical HMO in brain development and cognitive ability, was linked to weight gain, height gain, head circumference, and body composition, which highlights the significance of this HMO, which we posit confers physical growth in a secretor-specific manner alongside other milk components [30,31,43,47,48,54]. The second HMO of interest, 3’SL, had contradictory findings on weight gain, with positive associations in studies conducted by [18,30,38,43], and negative associations in studies conducted by [31,32]. 3’SL was additionally linked to height gain, head growth, and body composition [31,38,39,43,48,51,54]. The reason for this inconsistency in weight gain could be due to microbiota or macronutrients such as lipids and proteins found in milk having a direct relationship with growth. This can be further understood in the context of high-income overnutrition, which manifests as pediatric obesity, notably when breastfeeding is supplemented with formula containing 2’FL in mixed feeding settings occasioning over dosage on a critical HMO [35,39,54].

In our pooled analyses, LNFP II, a fucosylated HMO that was predominant in non-secretor mothers, was protective against excessive weight gain regardless of maternal secretor status, implying that early exposure to LNFP-II could offset early obesity risk, which is important to policymakers as a mitigation strategy in communities with paediatric obesity challenges [35]. With the microbiome featuring as an important indirect regulator of infant growth, we explore its role in this early critical event, fully aware of the consequences of dysbiosis induced by antibiotics [74]. Breastfed infants appear to have better outcomes because breast milk provides both HMOs and commensal bacteria (microbiota) in early life, with the former acting as prebiotics that are metabolized by the latter and thus playing a critical role in immune maturation and the latter enhancing infant health by preventing pathogen adhesion and promoting gut colonization by beneficial microbes [6,79]. Because milk oligosaccharides are structurally similar to intestinal mucin O-glycans, bacterial glycosidases digest carbohydrates from the intestine’s protective mucin layer, allowing pathobionts such as Enterobacteriaceae to cross-feed [80]. Any factor that can lead to microbiota dysbiosis early in life — such as inadequate breastfeeding, prolonged diarrhea, or antibiotics which eliminates crucial taxa for preserving homeostasis, leaving vacancies to be filled by blooms of pathobionts that may be more effective at extracting energy, predisposing some infants to obesity later in life [74,81]. Furthermore, HMOs function as receptor decoys, preventing bacterial adhesion to mucosal surfaces and thus preventing diarrhea [13]. They also reduce the pH of the infant’s digestive tract, increasing the proportion of beneficial bacteria *B. longum* while decreasing *E. coli* and *C. perfringens* [82]. In this context, we argue that children who are less sick will grow faster.

### Strengths and limitations

The strength of this study was its ability to summarize and quantify evidence for the role of HMOs on three interconnected early growth metrics, namely brain development, physical growth, and cognitive development, using clearly defined research question. We were cognizant of the fact evidence synthesis requires adequate representation for validity of the findings. We also included a wide spectrum of systematically selected studies from high and low income countries which helps to understand the influence of HMOs in regions of the world with highest undernutrition burden and associated highest neonatal and infant mortality rates globally which is critical for decision-making.

The main limitation of the studies was inconsistent reporting of HMO concentration; some authors used mean and standard deviation while others used median and IQR, which had to be converted, potentially losing some accuracy. Second, the random-effects model we used to calculate the pooled effect reports four heterogeneity statistics by default, which makes deciding what to interpret and what to leave out challenging. In this case, we reported both the τ^2^ statistic, which is insensitive to changes in the number of studies and their precision but is often difficult to interpret, and the I^2^ statistic, which is insensitive to changes in the number of studies and heavily depends on the precision of the included studies but is easier to interpret the percentage. Finally, we had only 12 comparing HMO concentrations between secretors and non-secretors.

### Conclusion and future opportunities

This study revealed that early-life exposure to 2’FL and 3’SL, which are abundant in secretor mothers, confer early childhood growth advantage to breastfeeding infants via optimal brain development and thus cognitive development. The proportion of secretor mothers in our study was 78%, implying that 22% of infants on non-secretors have no access to 2’FL and only have sub-optimal access to critical 3’SL via breastfeeding, both of which play critical roles in early childhood growth and development. Key emerging issues warrant a further investigation in future studies. There is a need for well-designed, sufficiently powered studies that distinguish between secretor and non-secretor women. Since suboptimal breastfeeding or poor maternal nutrition is linked poor developmental outcomes, it is important that these infants are supplemented with key HMOs that are linked with optimal growth. Unlike in the past, when this seemed impossible, it is now possible due to technological advances that have enabled large-scale production of key HMOs such as 2’FL, which is registered as a safe ingredient in infant formula in Europe and the USA. On the other hand, there is a risk of pediatric obesity, which may persist into adulthood. The finding that LNFP II is protective against pediatric obesity is of great interest, and more research is needed to confirm the findings. If successful, supplementation with this HMO holds the promise of new therapeutic applications to address obesity, a global concern. If successful, this HMO supplementation holds the promise of new therapeutic applications to address obesity, which is a global concern.

## Supporting information

PRISMA checklist

## Abbreviations

2’FL: 2-fucosyllactose
3FL: 3-fucosyllactose
DFLac: difucosyllactose
DFLNH: difucosyllacto-N-hexaose
DFLNT: difucosyllacto-N-tetraose
FLNH: fucosyllacto-N-hexaose
LNFP I: lacto-N-fucopentaose I
LNFP II: lacto-N-fucopentaose II
LNFP III: lacto-N-fucopentaose III
3’SL: 3-sialyllactose
6’SL: 6-sialyllactose
DSLNT: disialyllacto-N-tetraose
DSLNH: disialyllacto-N-hexaose
LST b: sialyllacto-N-tetraose b
LST c: sialyllacto-N-tetraose c
FDSLNH: fucodisialyllacto-N-hexaose
LNH: lacto-N-hexaose
LNnT: lacto-N-neotetraose
LNT: lacto-N-tetraose

## Author Contributions

Conceptualization: MM, RN and DW

Methodology: MM and HK

Investigation: MM, HK, LO and HA

Statistical analysis: MM and HA

Writing – original draft: MM and HA

Writing – review & editing: MM, RN and DW

## Data Availability Statement

All the data included was extracted from the selected original articles. The code used for the meta-analysis can be requested from the corresponding author.

## Supplementary legends

**Supplementary Table 1: PubMed search syntax for the following research question:** What effect do HMOs in human milk have on brain development, infant growth velocity, and neurodevelopment in breastfed infants?

**Supplementary table 2. Newcastle-Ottawa Scale (NOS) for Assessing the Quality of cross-sectional and cohort studies.** This tool measures four domains including: participant selection, comparability, exposure, and outcome. The scoring is based on the number of stars, with longitudinal studies receiving up to nine stars. Based on the stars received, a study can be rated high, moderate or low.

**Supplementary Table 3:** The effect of removal of an influential study on overall pooled estimate and heterogeneity statistics.

