## Supplementary Files for "The role of maternal secretor status and human milk oligosaccharides on early childhood development: A systematic review and meta-analysis"

**Supplementary Table 1: Pubmed Search syntax for the research question:** What effect do HMOs in human milk have on brain development, infant growth velocity, and neurodevelopment in breastfed infants?

| **PubMed** |
|---|
| ("breastfeeding"[MeSH Terms] OR ("breast"[All Fields] AND "feeding"[All Fields]) OR "breast feeding"[All Fields] OR "breastfeeding"[All Fields] OR "breastfeedings"[All Fields] OR "breastfeeders"[All Fields]) AND ((("milk, human"[MeSH Terms] OR ("milk"[All Fields] AND "human"[All Fields]) OR "human milk"[All Fields] OR ("human"[All Fields] AND "milk"[All Fields])) AND "oligosaccharide"[All Fields]) OR (("milk, human"[MeSH Terms] OR ("milk"[All Fields] AND "human"[All Fields]) OR "human milk"[All Fields] OR ("human"[All Fields] AND "milk"[All Fields])) AND "sugar"[All Fields])) AND ((("infant"[MeSH Terms] OR "infant"[All Fields] OR "infants"[All Fields] OR "infant s"[All Fields]) AND ("growth and development"[MeSH Subheading] OR ("growth"[All Fields] AND "development"[All Fields]) OR "growth and development"[All Fields] OR "growth"[All Fields] OR "growth"[MeSH Terms] OR "growths"[All Fields])) OR ("no to hattatsu"[Journal] OR "brain dev"[Journal] OR ("brain"[All Fields] AND "development"[All Fields]) OR "brain development"[All Fields]) OR "neurodevelopment"[All Fields] OR ("cogn dev"[Journal] OR ("cognitive"[All Fields] AND "development"[All Fields]) OR "cognitive development"[All Fields])) |

All searches were confined to English-language studies conducted between 2000 and June 30th 2023.

**Supplementary Table 2:** Newcastle-Ottawa Scale (NOS) for assessing the quality of cohort and cross-sectional studies

| # | Author | Selection | Comparability | Outcome | GRADE rating |
|---|---|---|---|---|---|
| 1 | Alderete et al., 2015 | ★★★ | ★ | ★★ | medium |
| 2 | Berger et al., 2020 | ★★★ | ★★ | ★★★ | high |
| 3 | Berger et al., 2020b | ★★ | ★★ | ★★★ | medium |
| 4 | Berger et al., 2022 | ★★ | ★★ | ★★★ | medium |
| 5 | Binia et al., 2021 | ★★★★ | ★★ | ★★★ | high |
| 6 | Charbonneau et al., 2016 | ★★★★ | ★★ | ★★★ | high |
| 7 | Cheema et al., 2022 | ★★ | ★★ | ★★★ | medium |
| 8 | Cho et al., 2021 | ★★★ | ★★ | ★★ | medium |
| 9 | Davis et al., 2017 | ★★ | ★★ | ★★ | medium |
| 10 | Dou et al., 2023 | ★★ | ★★ | ★★ | medium |
| 11 | Ferreira et al., 2021 | ★★★ | ★★ | ★★ | medium |
| 12 | Lagström et al., 2020 | ★★★★ | ★★ | ★★★ | high |
| 13 | Larsson et al., 2019 | ★★★ | ★★ | ★★ | medium |
| 14 | Mansell et al., 2023 | ★★ | ★★ | ★★★ | high |
| 15 | Menzel et al., 2021 | ★★★ | ★★ | ★★★ | high |
| 16 | Nuzhat et al., 2022 | ★★ | ★★ | ★★ | medium |
| 17 | Oliveros et al., 2021 | ★★ | ★★ | ★★ | low |
| 18 | Rozé et al., 2022 | ★★★ | ★★ | ★★ | medium |
| 19 | Saben et al., 2021 | ★★ | ★★ | ★★ | medium |
| 20 | Samuel et al., 2022 | ★★ | ★★ | ★★ | medium |
| 21 | Sprenger et al., 2017 | ★★★ | ★★ | ★★ | medium |
| 22 | Tonon et al., 2019 | ★★ | ★★ | ★★ | medium |
| 23 | Wang et al., 2020 | ★★★ | ★★ | ★★★ | medium |
| 24 | Willemsen et al., 2023 | ★★★ | ★★ | ★★★ | medium |

**GRADE** = Grading of Recommendations Assessment, Development and Evaluation

**Supplementary Table 3:** The effect of removal of an influential study on overall pooled estimate and heterogeneity statistics

| Analysis | n | g (95% CI) | p value | $\tau^2$ (95% CI) | p-value |
|---|---|---|---|---|---|
| **2'FL** analysis with Influential publication | 13 | 3.84 (1.54; 6.14) | 0.003 | 13.80 (7.04; 40.27) | < .001 |
| **2'FL** analysis without Influential publication | 12 | 2.80 (2.40; 3.20) | < .001 | 0.28 (0.09; 1.07) | < .001 |
| **3' SL** analysis with Influential publication | 10 | 1.21 (-1.16; 3.57) | 0.277 | 10.63 (4.97; 36.78) | < .001 |
| **3'SL** analysis without Influential publication | 9 | 0.21 (-0.16; 0.59) | 0.230 | 0.19 (0.05; 0.77) | < .001 |

n = Number of studies; $\tau^2$ = heterogeneity variance (tau) statistic which quantifies the variance of the true effect sizes underlying our data, if it does not include zero then there is heterogeneity in the data, $\tau^2$ was estimated using the restricted maximum likelihood (REML) estimator. The confidence interval (CI) around the pooled effect was computed using Knapp-Hartung adjustments. The study by Saben et al 2021, was a significant outlier that influenced the pooled statistic. Its exclusion reduced the $\tau^2$ statistics significantly while only moderately reducing the pooled estimates for both 2'FL and 3'SL.
